# The impact of co-circulating pathogens on SARS-CoV-2/COVID-19 surveillance: How concurrent epidemics may decrease true SARS-CoV-2 percent positivity

**DOI:** 10.1101/2021.06.08.21258533

**Authors:** Aleksandra Kovacevic, Rosalind M. Eggo, Marc Baguelin, Matthieu Domenech de Cellès, Lulla Opatowski

## Abstract

**Background:** Circulation of non-SARS-CoV-2 respiratory viruses during the COVID-19 pandemic may alter quality of COVID-19 surveillance, with possible consequences for real-time analysis and delay in implementation of control measures. Here, we assess the impact of an increased circulation of other respiratory viruses on the monitoring of positivity rates of SARS-CoV-2 and interpretation of surveillance data.

**Methods:** Using a multi-pathogen Susceptible-Exposed-Infectious-Recovered (SEIR) transmission model formalizing co-circulation of SARS-CoV-2 and another respiratory we assess how an outbreak of secondary virus may inflate the number of SARS-CoV-2 tests and affect the interpretation of COVID-19 surveillance data. Using simulation, we assess to what extent the use of multiplex PCR tests on a subsample of symptomatic individuals can support correction of the observed SARS-CoV-2 percent positive during other virus outbreaks and improve surveillance quality.

**Results:** Model simulations demonstrated that a non-SARS-CoV-2 epidemic creates an artificial decrease in the observed percent positivity of SARS-CoV-2, with stronger effect during the growth phase, until the peak is reached. We estimate that performing one multiplex test for every 1,000 COVID-19 tests on symptomatic individuals could be sufficient to maintain surveillance of other respiratory viruses in the population and correct the observed SARS-CoV-2 percent positive.

**Conclusions:** This study highlights that co-circulating respiratory viruses can disrupt SARS-CoV-2 surveillance. Correction of the positivity rate can be achieved by using multiplex PCR, and a low number of samples is sufficient to avoid bias in SARS-CoV-2 surveillance.

**Summary:** COVID-19 surveillance indicators may be impacted by increased co-circulation of other respiratory viruses delaying control measure implementation. Continued surveillance through multiplex PCR testing in a subsample of the symptomatic population may play a role in fixing this problem.

## Introduction

Severe acute respiratory syndrome coronavirus 2 (SARS-CoV-2) caused a worldwide pandemic of coronavirus disease 2019 (COVID-19) prompting the need for global disease surveillance. Such global surveillance aims to monitor trends in COVID-19 to identify patterns of transmission and progression, estimate morbidity and mortality, and assess the impact of control measures [1]. Implementation of community control strategies (i.e., mask wearing, lockdowns, social distancing, and school closures) to limit COVID-19 transmission also impacted other common respiratory viruses, causing a drop in their detection. Indeed, national and international lockdowns and travel restrictions in March 2020 caused a sharp decline in seasonal influenza circulation in the United States, while the measures almost completely eliminated influenza in Southern Hemisphere countries such as Australia, Chile, and South Africa during their typical influenza season, June-August 2020 [2]. A similar drop in detection was observed for respiratory syncytial virus (RSV) [3, 4], while rhinovirus activity appeared to be low during the lockdown period [5]. However, when these measures are relaxed, circulation of viruses can reoccur, mediated by the resumption of social interactions. For example, data from NSW, Australia showed a surge in rhinovirus once schools reopened in mid-May 2020 [5]. Similarly, in Hong Kong, in England, and in France, school reopening in the fall of 2020 coincided with increased rhinovirus activity, particularly in school-aged children [6, 7, 8].

While the role of children in the transmission of SARS-CoV-2 is still debated and children are thought to be less efficient in transmitting the virus than adults, children are frequently the main drivers of transmission of other respiratory viruses such as rhinovirus and influenza [9-13]. Indeed, the sharp increase in rhinovirus detection among adults admitted to hospitals observed in Southampton, UK followed the reopening of schools in September 2020 [14]. As a consequence, circulation of other respiratory viruses during the pandemic may have an impact on COVID-19 surveillance. SARS-CoV-2 can cause a wide range of symptoms varying in severity [15], and many of these resemble symptoms of other influenza-like illnesses (ILIs) and acute respiratory infections (ARIs) including influenza viruses, RSV, rhinovirus, and others. For example, the rise in symptomatic cases in weeks 34-40 observed in Canada reflected rapidly rising enterovirus/rhinovirus disease activity rather than COVID-19 [16]. Delay in trend interpretation may lead to delayed decision making and control measure implementation, which may have substantial negative consequences on the public health system due to the exponential nature of COVID-19 epidemic [17].

The World Health Organization recommends considering test positivity proportion over a two-week period as a key epidemiological indicator to assess and classify the level of community transmission [18], and therefore, other respiratory viruses could generate misinterpretation of COVID-19 surveillance data. For instance, Public Health France reports showed a decrease in SARS-CoV-2 symptomatic percent positive in September 2020, which is the time when schools reopened in France, although the daily number of symptomatic COVID-19 cases did not decrease [19]. We hypothesized that this marked decrease in SARS-CoV-2 symptomatic percent positive could be caused by increased co-circulation of another respiratory virus.

In this study, we used mathematical modelling to investigate the consequences of an increased circulation of other respiratory viruses both in terms of the number of SARS-CoV-2 tests taken, but also in the monitoring of percent positivity of SARS-CoV-2 tests and interpretation of surveillance data.

## Methods

### SARS-CoV-2 surveillance data

We focus here on the surveillance of symptomatic individuals as they are more likely to request SARS-CoV-2 tests. We used data on SARS-CoV-2 tests and positive cases in metropolitan France from Public Health France (SI-DEP) for the period from 08/24/20 to 19/10/20 [20]. Symptomatic individuals who started presenting symptoms 0-4 days before PCR test were classified as symptomatic (and referred throughout as symptomatic). Asymptomatic individuals, individuals who started presenting symptoms 5-14 days or more before the COVID-19 test, and individuals whose symptomatic status was not recorded, were classified as other. We separated the data in such way because individuals having recent symptoms relative to the time of testing were more likely to be infected with a non-SARS-CoV-2 respiratory virus with shorter symptom duration where symptoms typically peak at 1-3 days [21], as opposed to individuals who were presenting symptoms for a long time before requesting COVID-19 test.

### Hospitalization data

COVID-19 hospitalization data were acquired from Public Health France, SI-VIC (Information system for monitoring victims) from 08/24/20 to 19/10/20 and daily numbers were aggregated [22].

### Neutral transmission model

We developed a multi-pathogen Susceptible-Exposed-Infectious-Recovered (SEIR) transmission model to explore and illustrate how cocirculation of another respiratory virus (called virus-2) with symptomatic overlap during the ongoing COVID-19 pandemic may affect test percent positivity. We build upon a model of two circulating pathogens [23] by adding Exposed compartment for both viruses to more closely match the natural history of SARS-CoV-2 and other respiratory viruses (Figure 2). The model is neutral assuming no interaction between two pathogens, i.e., no change in infectiousness of co-infected classes and no change in the probability of acquisition of a second infection following the first infection (sections S1.1 – S1.4, Supplementary Material).

### Testing model

We modelled the total number of tests in symptomatic individuals by considering four different reasons for testing symptomatic individuals: *(i)* symptomatic individuals infected with SARS-CoV-2 (*T*_*COV*_); *(ii)* we assumed that contact tracing and testing on a day *t* would generate an increase in test demand after some contact tracing delay, *d* in days, where we considered only contacts presenting symptoms at the time of testing (*T*_*CONTACTS*_); *(iii)* symptomatic individuals infected with virus-2 (*T*_*V2*_); and finally *(iv)* we also considered a baseline number of symptomatic tests (*T*_*b*_), i.e., symptomatic individuals getting tested for SARS-CoV-2, but truly negative for SARS-CoV-2 and negative for virus-2.

Thus, the total number of symptomatic SARS-CoV-2 tests *T* on a given day *t* is given by:

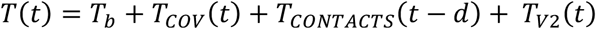

Observed SARS-CoV-2 symptomatic percent positive *P*_*+*_ on a day *t* is given by:

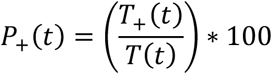

where *T*_*+*_ corresponds to the number of SARS-CoV-2 positive tests among symptomatic individuals changing in time (see S2.1, Supplementary Material for details).

### Testing strategies

To investigate if it was possible to correct the observed SARS-CoV-2 symptomatic positivity rate using the results of tests for virus-2 in a subsample of the population, we modelled the use of multiplex PCR tests along with standard PCR SARS-CoV-2 tests on a subsample of symptomatic individuals. We tested if results of multiplex tests could be used to maintain the surveillance of other viruses and determined what correction of the observed SARS-CoV-2 percent positive was needed during other virus outbreaks.

Therefore, we introduced two different types of tests into the model: RT-PCR (reverse transcription-polymerase chain reaction) SARS-CoV-2 test with sensitivity *s*_*pcr*_ and a multiplex PCR test that simultaneously tests for multiple pathogens, with average sensitivity *s*_*m*_ to detect SARS-CoV-2 or virus-2.

We assumed that the proportion of multiplex tests actually carried out across all tests on a given week was *m*.

We estimated the corrected SARS-CoV-2 symptomatic percent positive *P*_*C+*_ on a day *t* was computed as:

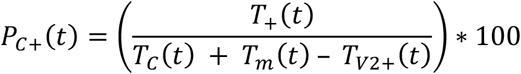

where *T*_*C*_*(t)* represents the corrected total number of symptomatic SARS-CoV-2 tests on a given day *t, T*_*m*_*(t)* is the total number of symptomatic multiplex tests on a given day *t* and is given by:

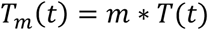

and *T*_*V2+*_ is the number of confirmed positive symptomatic virus-2 cases detected with multiplex tests:

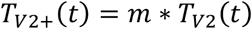

The corrected total number of symptomatic SARS-CoV-2 tests *T*_*C*_*(t)* on a given day *t* is:

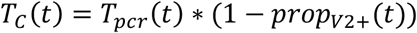

where *T*_*pcr*_*(t)* represents the number of symptomatic SARS-CoV-2 tests on day *t* and *prop*_*V2+*_*(t)* is the proportion of symptomatic virus-2 confirmed positive cases among all multiplex tests (S2.2, Supplementary Material).

To account for the impact of imperfect sensitivity of tests and multiplex test sample size in the uncertainty of indicators, we added a stochastic observation model. The observed number of positive tests for SARS-CoV-2 (*T*_*COV*_) and virus-2 (*T*_*V2+*_) respectively, were calculated assuming a binomial distribution (S2.3, Supplementary Material).

### Simulation study

We simulated the introduction of an outbreak of virus-2 in a population with circulating SARS-CoV-2 coinciding with a particular event that increases social interactions (e.g., school reopening). We assumed 6% of the population had already been infected and became immune to SARS-CoV-2, which matches the situation of France in September 2020 at the time of school reopening [24]. We also assumed that 70% of the population is either immune to virus-2 or will not get exposed to the virus at all due to generally never being in contact with school children, social distancing measures, and implemented community control strategies (S1.5, Supplementary Material). We tracked two key epidemiological indicators: number of tests requested and percent positivity.

### Sensitivity analyses

First, we varied the sensitivity of multiplex PCR test sensitivity (*s*_*m*_), while maintaining the same sensitivity of SARS-CoV-2 PCR test (*s*_*pcr*_) to investigate its impact on the corrected percent positive of SARS-CoV-2. We also evaluated a range of proportion values of multiplex tests (*m* = 0.0005; 0.001; 0.002; 0.005) to assess how sample size of multiplex tests affects the correction of the observed symptomatic SARS-CoV-2 percent positive. Then, we conducted additional sensitivity analyses varying *s*_*pcr*_ and the proportion of symptomatic individuals infected with virus-2 (*s*_*2*_), while maintaining other parameters the same. Finally, we investigated the impact of R_0_ (basic reproduction number) of virus-2 and the initial proportion of the population immune to SARS-CoV-2 and to virus-2.

R (version 4.0.3) and RStudio (version 1.3.1093) were used for modelling transmission, testing, and all statistical analyses [25, 26].

## Results

Data from Public Health France show a decrease in symptomatic percent positive for SARS-CoV-2 in France after September 1^st^, 2020, synchronous with school reopening, while the hospitalization data show a steady increase (Figure 3D). However, this decrease in positivity rate was not due to a decline in SARS-CoV-2 positive tests among symptomatic individuals (Figure 1a). Moreover, the number of tests for symptomatic individuals increased during September 2020 as well as the number of other tests (Figure 1b).

**Figure 1.**
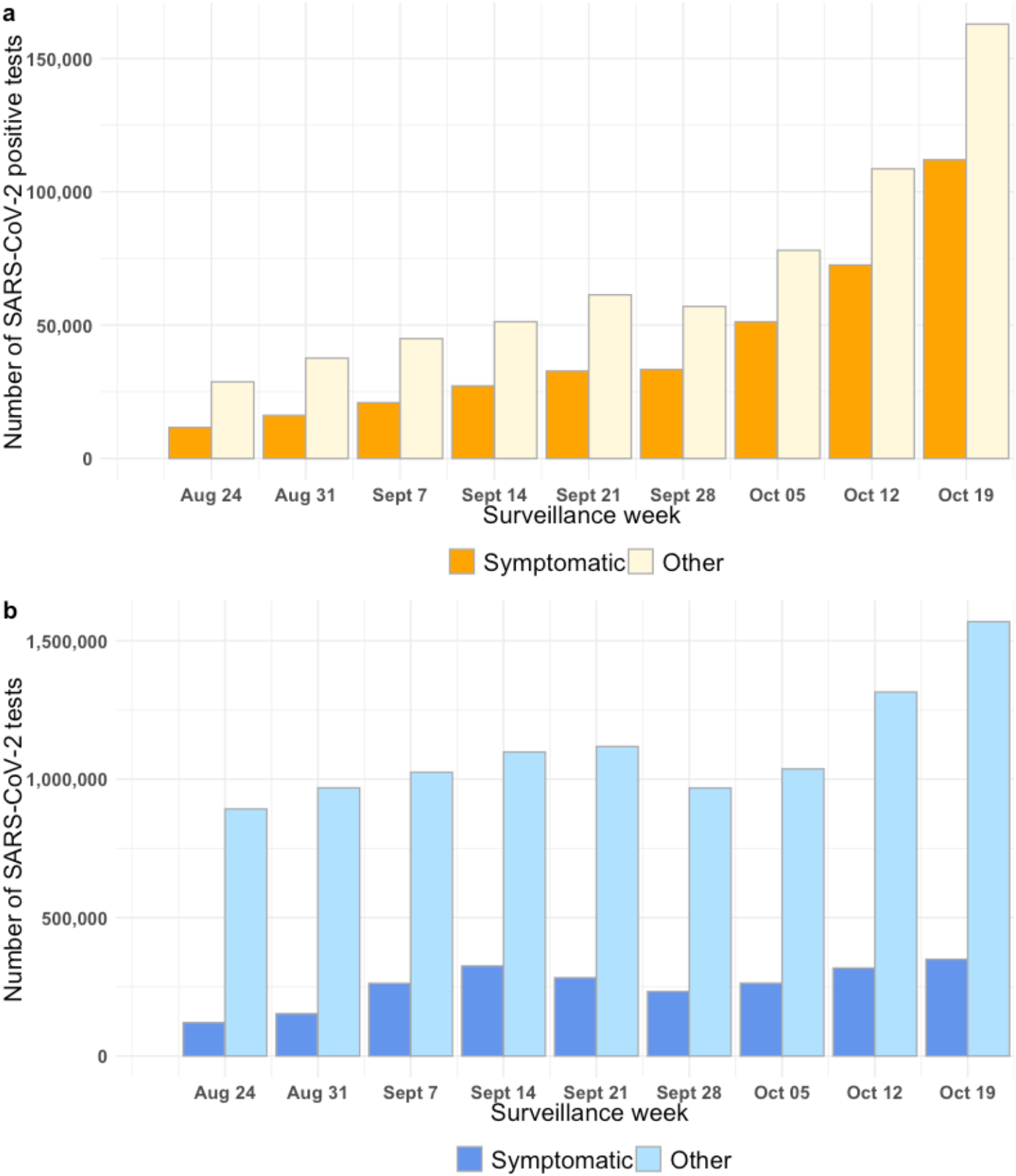
SARS-CoV-2 weekly testing data in France from 08/24/2020 to 10/19/2020 [20]. **a)** Number of positive SARS-CoV-2 tests **b)** Total number of SARS-CoV-2 tests.

**Figure 2.**
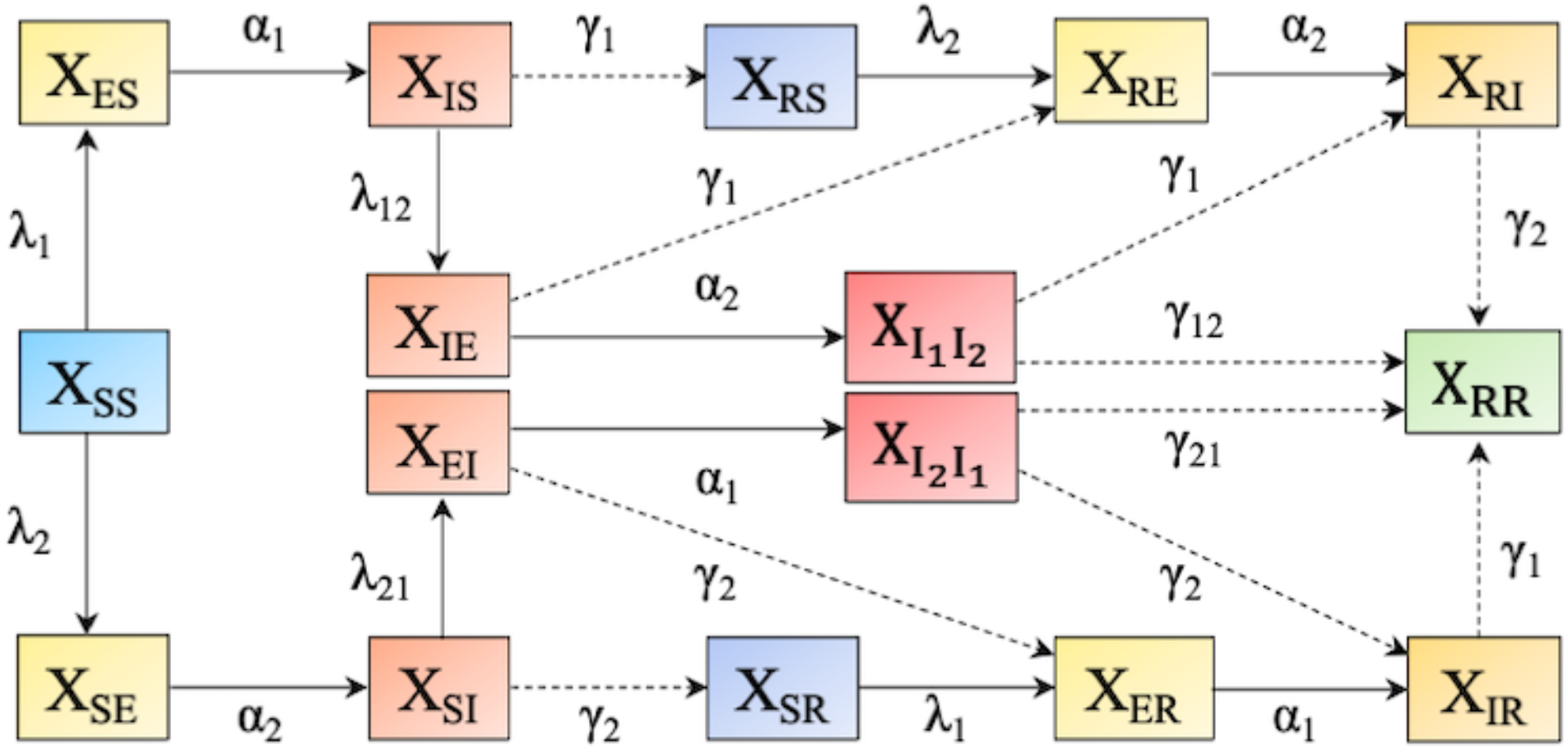
Model formalizing the transmission of two cocirculating viruses in a population. Individuals are split into compartments (*X*_*ij*_) according to their status with respect to the two pathogens. In *X*_*ij*_, subscript *i* specifies individual’s status to virus-1 (SARS-CoV-2) and subscript *j* status to virus-2. Letters S, E, I, and R stand for Susceptible, Exposed, Infected, and Recovered, respectively. Rates of transition between compartments are infectiousness onset rates (*α*_*1*,_ *α*_*2*_) and recovery rates (*γ*_*1*_, *γ*_*2*_). λ_1_ and λ_2_ stand for the forces of infection for susceptible hosts by virus-1, i.e., SARS-CoV-2 and virus-2, respectively; *λ*_*21*_ and *λ*_*12*_ are forces of infection by virus-1 (*λ*_*21*_) and virus-2 (*λ*_*12*_) for hosts already infected by the other pathogen (S1.3, Supplementary Material).

**Figure 3.**
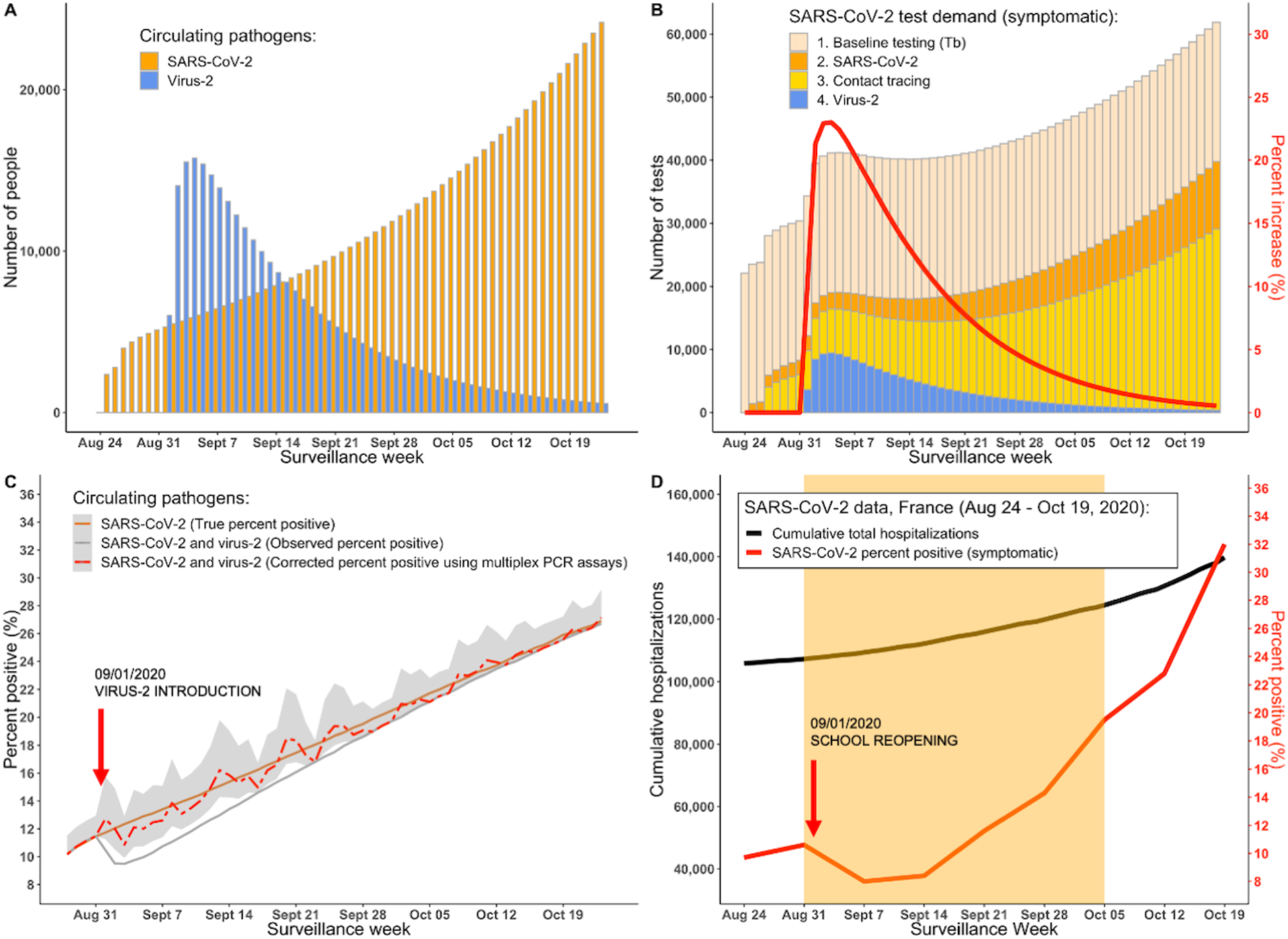
Impact on surveillance indicators - Model simulations of the virus-2 introduction in a population with the ongoing SARS-CoV-2 epidemic and COVID-19 hospitalization and testing data in France from 08/24/2020 to 10/19/2020. **A**. Simulations of true daily symptomatic incidences of SARS-CoV-2 and virus-2. **B**. Simulations of SARS-CoV-2 test demand (symptomatic) including tests requested by individuals infected with both viruses, contacts of previously identified cases, and a constant testing baseline. Red line represents percent increase in SARS-CoV-2 test demand. **C**. Simulated SARS-CoV-2 percent positive among all tests for symptomatic individuals. 09/01/2020 marks the introduction time of virus-2 in the population (e.g., following the school reopening). The outbreak of virus-2 decreases the observed COVID-19 percent positive (grey solid line) relative to the true percent positive (orange solid line) assuming testing demand and surveillance is not affected by virus-2. Observed percent positive can be corrected by testing a proportion (*m*) of symptomatic cases with multiplex PCR assays. Corrected percent positive is conducted with testing frequency of 1 multiplex per 1,000 daily tests (red dashed line with 95% confidence intervals). All simulations are run assuming *s*_*pcr*_ = 95% and *s*_*m*_ = 90% (S1.5, Supplementary Material). **D**. SARS-CoV-2 hospitalizations [22] and symptomatic percent positivity [20] in France from 08/24/20 to 10/19/20. Orange shaded area represents observed underestimation of SARS-CoV-2 symptomatic percent positivity.

In our simulation study, the outbreak of virus-2 lasted for 1.5 months with a peak reached after two weeks (Figure 3A). These first weeks of virus-2 circulation led to up to 23% increase in the number of daily tests performed on symptomatic individuals (Figure 3B). The observed SARS-CoV-2 percent positive among symptomatic tests decreased sharply in the first two weeks, underestimating the true percent positivity by up to 3% (23% relative decrease), and then progressively increased to converge to the true percent positivity when the outbreak was nearly extinct (Figure 3C). We determined that a testing frequency *m* of 0.1% (m=0.001; i.e., performing one multiplex tests for every 1,000 symptomatic tests), which most closely represents realistic proportion of multiplex tests currently used in France to test for non-SARS-CoV-2 respiratory viruses [7], was sufficient to provide a correction that closely follows real percent positive of SARS-CoV-2 (Figure 3C).

Sensitivity analyses showed that changing the sensitivity of the multiplex assays (*s*_*m*_) affected the corrected percent positive of SARS-CoV-2, with higher values of *s*_*m*_ providing better estimates for real percent positive (S2.4.1, Supplementary Material). Similarly, the correction of the observed SARS-CoV-2 percent positive was negatively affected by the lower proportion of the multiplex PCR testing (*m*) used in the model, and vice versa. Larger proportion of multiplex PCR tests used improved the correction of the observed SARS-CoV-2 percent positive (S2.4.2, Supplementary Material). The correction quality was not affected by varying *s*_*pcr*_, *s*_*2*_, *R*_*0*_, or the initial proportion of the population immune to SARS-CoV-2 and to virus-2. (S2.4.3 – S2.4.7, Supplementary Material).

## Discussion

The implementation of the community control strategies to limit transmission of SARS-CoV-2 also decreased the circulation of other respiratory viruses. However, once control measures are relaxed and when social interactions are resumed, we are likely to observe increased activity of other respiratory viruses [6, 7, 8]. Using model simulations of two co-circulating pathogens, we showed that an outbreak of a secondary respiratory virus during COVID-19 pandemic may increase SARS-CoV-2 testing demand and, as a consequence, hinder the detection of the initial increase of the true percent positivity of SARS-CoV-2 and may lead to the overall underestimation of the SARS-CoV-2 epidemic. We proposed to correct the observed positivity rate of SARS-CoV-2 by using multiplex PCR testing in a subsample of the symptomatic population. We estimate that performing one multiplex test for every 1,000 of COVID-19 tests could be enough to significantly improve real time epidemiological interpretation.

Underreporting of infection is a challenge in all pandemics and epidemics including this one [27, 28], and we show here that short-term alterations in surveillance due to other epidemics may affect interpretation of COVID-19 trends. Indeed, after schools reopened, multiplex testing detected circulation of rhinovirus in France (S2.5, Supplementary Material), which supports our hypothesis that secondary virus might have been responsible for decreased COVID-19 epidemic indicators. Positivity rate, a key indicator monitored for epidemic control and public health decision-making [18] appears particularly sensitive to this effect. Moreover, maintaining surveillance of other respiratory viruses should aid public health officials to better anticipate increases in SARS-CoV-2 testing demand.

The use of multiplex PCR assays for SARS-CoV-2 detection in a sample of symptomatic cases has already proved effective in detecting other respiratory viruses: a study in Northern California in March 2020 found that 26.7% of symptomatic patients who tested negative for SARS-CoV-2 were positive for one or more additional pathogens, most often rhinovirus/enterovirus [29]. Another study found that 13.1% of patients who tested negative for SARS-CoV-2 were positive for at least one non-SARS-CoV-2 respiratory viral pathogen [30]. These studies not only reflect that the non-SARS-CoV-2 respiratory viruses are circulating in our communities, but that symptomatic cases due to other respiratory pathogens contribute to the overall number of negative SARS-CoV-2 tests. Increase in the hospitalization data and the number of SARS-CoV-2 positive tests in symptomatic individuals during observed decrease in symptomatic percent positive suggest that decrease in COVID-19 cases was not responsible for the decrease in test positivity rate. Considering that rhinovirus can cause fever and severe sore throat in children, and epidemics of rhinovirus are common on school reopening [31, 32], it is likely that symptomatic children and parents requested testing for SARS-CoV-2, thus increasing the number of negative tests and inadvertently reducing the test-positivity rate during this time.

To keep our model simple, the mechanisms are a simplification of the real processes, and therefore, there are limitations. We did not incorporate testing capacity within this model, and we assumed that all symptomatic individuals requesting testing on a given day will be able to do so. Additionally, we assumed that interactions between viruses were neutral, meaning that the presence of one of the viruses did not affect (promote nor protect against) infection with the other virus. Recent studies have suggested some possible protection from COVID-19 infection conferred by rhinovirus interference with SARS-CoV-2 replication kinetics and this may warrant further exploration at the population level [33].

We proposed a method to correct the observed positivity rate of SARS-CoV-2 during an outbreak of another respiratory virus, to help reduce the overall underestimation of SARS-CoV-2 in the population. Clinical sensitivities between tests can differ markedly depending on the test manufacturer and in the case of multiplex PCR testing, sensitivity also depends on the pathogen being detected [30, 34]. With the overall high sensitivity of both SARS-CoV-2 and multiplex PCR tests, correcting the observed positivity rate could be a very effective way of minimizing the underestimation of the true COVID-19 burden in the community. Furthermore, multiplex testing, which in France is generally performed by ‘Sentinelles’ physicians on patients seen in the consultation to test for various respiratory viruses, could be incorporated into laboratory testing for SARS-CoV-2 to improve the surveillance and detection of other respiratory viruses, which in turn may improve detection of the real percent positive of SARS-CoV-2 individuals in the population.

Using modelling simulations, we highlight that co-circulating respiratory viruses impact COVID-19 surveillance. Our results demonstrate that systematic use of multiplex PCR tests on a subsample of symptomatic individuals is key to maintaining unbiased surveillance.

## Data Availability

N/A

## Acknowledgements

We want to thank David RM Smith for helpful discussion and valuable advice on multiplex testing, and Sylvie van der Werf, Cécile Souty, Pierre-Yves Boëlle, and Daniel Levy-Bruhl for their helpful discussion.

## Supplementary Material

### S1. Transmission model

#### S1.1 Model parameters

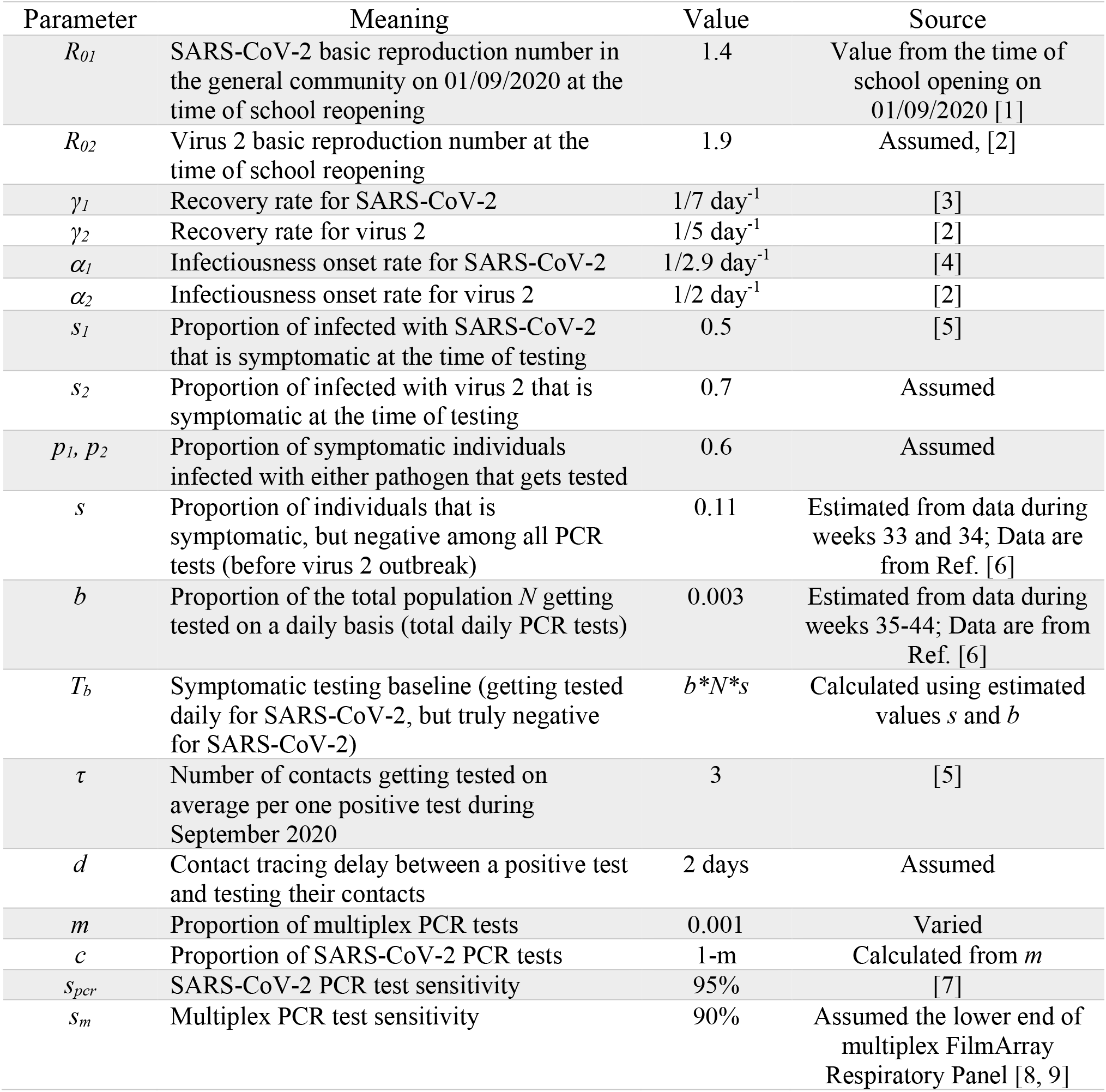

#### S1.2 Interaction parameters

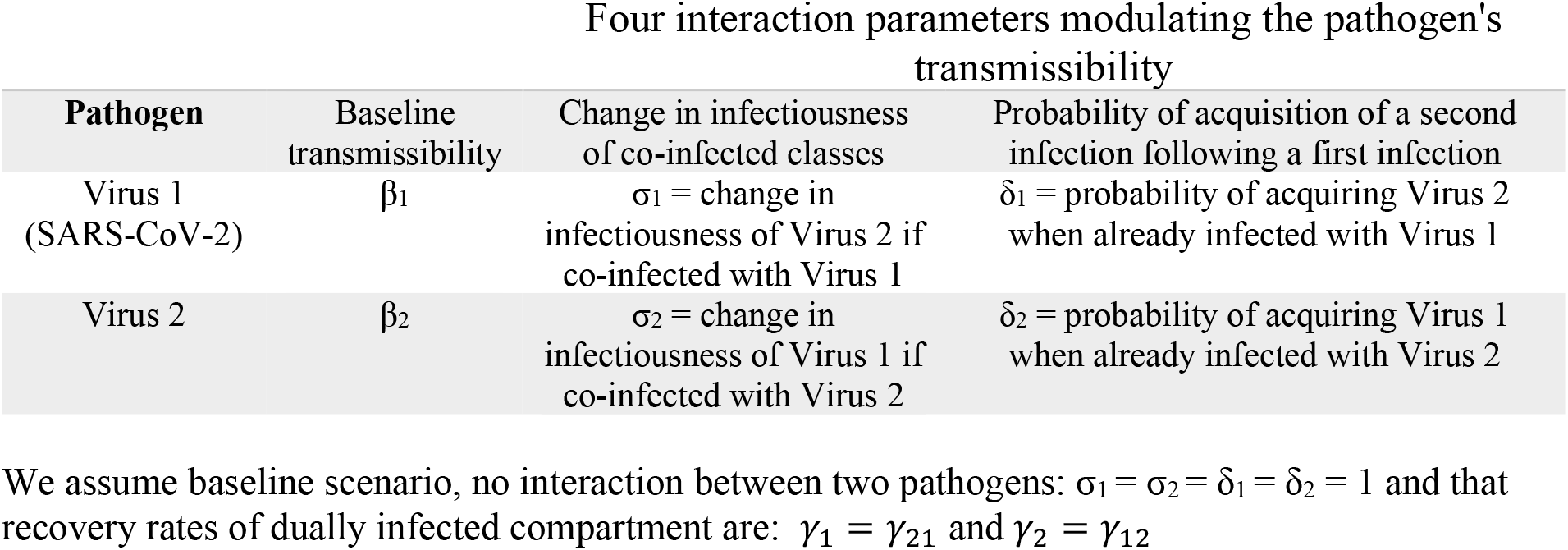

S1.2 Interaction parameters Four interaction parameters modulating the pathogen's transmissibility
| Pathogen | Baseline transmissibility | Change in infectiousness of co-infected classes | Probability of acquisition of a second infection following a first infection |
| --- | --- | --- | --- |
| Virus 1<br>(SARS-CoV-2) | $\beta_1$ | $\sigma_1$ = change in infectiousness of Virus 2 if co-infected with Virus 1 | $\delta_1$ = probability of acquiring Virus 2 when already infected with Virus 1 |
| Virus 2 | $\beta_2$ | $\sigma_2$ = change in infectiousness of Virus 1 if co-infected with Virus 2 | $\delta_2$ = probability of acquiring Virus 1 when already infected with Virus 2 |

We assume baseline scenario, no interaction between two pathogens: $\sigma_1 = \sigma_2 = \delta_1 = \delta_2 = 1$ and that recovery rates of dually infected compartment are: $\gamma_1 = \gamma_{21}$ and $\gamma_2 = \gamma_{12}$

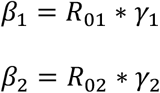

#### S1.3 Forces of infection

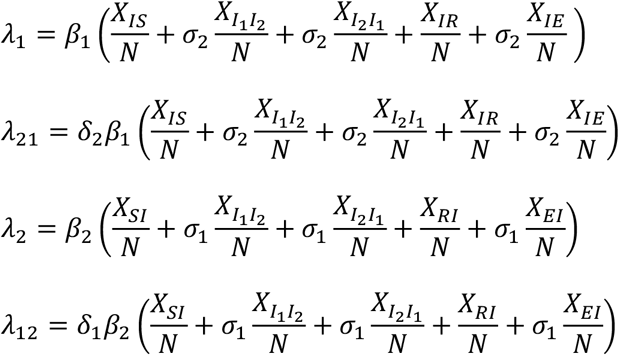

#### S1.4 Transmission model equations

Susceptible:

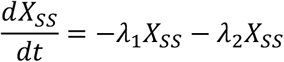

Exposed:

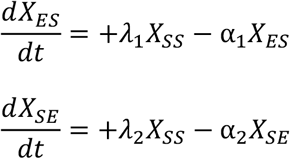

Infected:

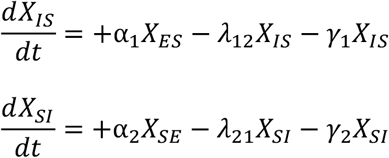

Infected with one virus, exposed to the other:

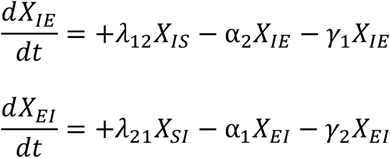

Double infected:

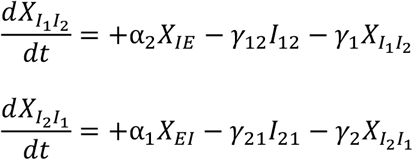

Recovered from one virus:

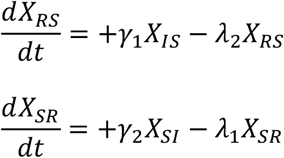

Recovered from one virus, exposed to the other:

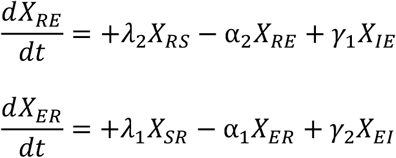

Recovered from one virus, infected with the other:

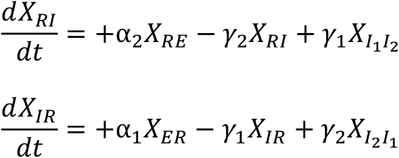

Recovered from both viruses:

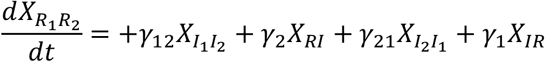

#### S1.5 Simulation study

##### Initialization

The modelling study was simulated on the population of 67 million people. Model was initialized assuming no interaction scenario, with 6% of the population being in *X*_*RS*_ compartment [10] and 70% of the population in *X*_*SR*_ compartment allowing for virus-2 outbreak to finish within two-month period. SARS-CoV-2 was initialized with 12,060 individuals in the Exposed (*X*_*ES*_) compartment and 42,210 individuals in the Infected (X_IS_) compartment from the week before. Virus-2 was introduced on day 7 with 300,000 individuals in total, with 270,000 being in the Infected (*X*_*SI*_) compartment and 30,000 individuals being in Exposed (*X*_*SE*_) compartment.

We simulated an outbreak of virus-2 for 8 weeks. Parameter values used in the study correspond to the real time values reported right before and during the time when a decrease in symptomatic percent positive of SARS-CoV-2 was observed in the French population – starting in the first week of September, the time of school opening. Two pathogens had different transmission characteristics at the time of an outbreak of virus-2 (SARS-CoV-2: R_01_ = 1.4, _*1*_ = 1/7 day^-1^, _*1*_ = 1/2.9 day^-1^; virus-2: R_02_ = 1.9, _*2*_ = 1/5 day^-1^, _*2*_ = 1/2 day^-1^) with virus-2 having a greater basic reproduction number (R_0_) because of its ability to spread easily among school children, and a faster recovery rate (i.e., shorter infectious period).

### S2. Testing model

#### S2.1 Number of tests

Baseline number of symptomatic tests (*T*_*b*_) is calculated by assuming that a constant proportion of population *N* is tested on a daily basis (*b*) with a proportion of this population being symptomatic (*s*) due to reasons other than COVID-19 infection or virus-2 outbreak. Baseline symptomatic tests are assumed to be negative for both SARS-CoV-2 and virus-2.

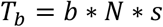

Number of tests due to symptomatic individuals infected with SARS-CoV-2 is calculated by assuming only a proportion of population infected with SARS-CoV-2 will present symptoms (*s*_*1*_) and request testing (*p*_*1*_).

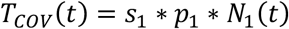

with *N*_*1*_ being the incident number of people infectious with SARS-CoV-2.

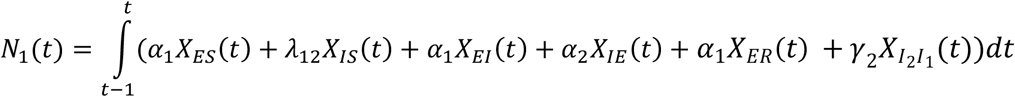

Similar approach was taken when calculating number of tests due to symptomatic individuals infected with virus 2 assuming that only a proportion of population infected with virus-2 will present symptoms (*s*_*2*_) and request testing (*p*_*2*_) for COVID-19.

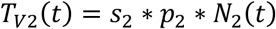

where *N*_*2*_ is the incident number of people infectious with virus-2, not including double infected individuals.

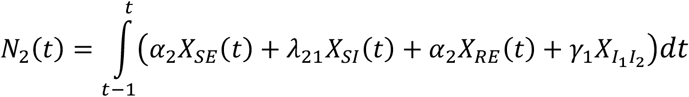

Double infected individuals are excluded from virus-2 count because ultimately, they will be counted towards positive COVID-19 cases and will not be excluded from the corrected total of daily tests when we conduct correction of the percent positive.

The number of symptomatic tests generated by contact tracing and testing around confirmed infected cases with SARS-CoV-2 on a day *t*, assuming a contact tracing delay *d* between a positive case and testing their contacts, is calculated as following:

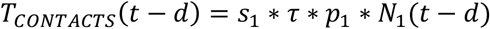

where *τ* represents number of contacts tested per one positive case, on average. We considered contacts of both symptomatic and asymptomatic confirmed index cases but only included symptomatic contacts in the total number of symptomatic tests. Among these symptomatic tests generated by contact tracing and testing around confirmed infected cases, the proportion of truly infected with SARS-CoV-2 is assumed to scale with the prevalence of SARS-CoV-2 in the population at the date of the test.

Prevalence of SARS-CoV-2 in the population *prev(t)* at time *t* is the proportion of population infected with SARS-CoV-2 at time *t*, it can be estimated by:

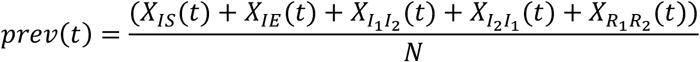

Number of SARS-CoV-2 positive tests among symptomatic individuals *T*_*+*_ on a given day *t* is calculated according to the following equation:

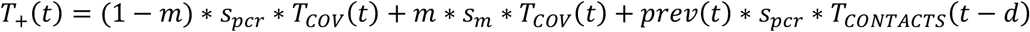

where *m* represents the proportion of multiplex PCR tests among total tests conducted on symptomatic individuals, *s*_*pcr*_ and *s*_*m*_ represent sensitivity of PCR SARS-CoV-2 test and PCR multiplex tests, respectively, with *prev* representing prevalence of SARS-CoV-2 in the population on a given day *t*.

#### S2.2 Correction of the observed SARS-CoV-2 symptomatic percent positive

We proposed a corrected version of the symptomatic percent positive of SARS-CoV-2 based on the results of multiplex PCR testing:

The proportion of symptomatic virus-2 confirmed positive cases among all multiplex PCR tests (*T*_*m*_) is given by:

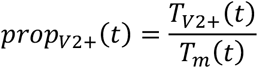

where *T*_*V2*_ is the confirmed positive symptomatic virus 2 cases and *T*_*m*_ is the number of symptomatic multiplex PCR tests.

Total symptomatic daily tests *T* on a given day *t* are sum of tests conducted with PCR COVID-19 tests:

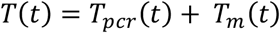

where *T*_*pcr*_*(t)* represents the number of symptomatic PCR SARS-CoV-2 tests on day *t*:

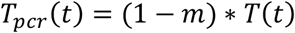

*T*_*m*_*(t)* is the total number of symptomatic multiplex tests on a given day *t* and is given by:

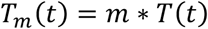

#### S2.3 Stochastic reporting of test results

The observed number of positive tests for SARS-CoV-2 (*T*_*COV*_) and virus-2 (*T*_*V2+*_) respectively, were modelled as follows:

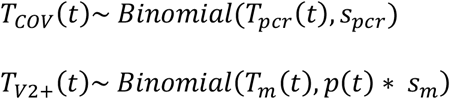

where *s*_*pcr*_ and *s*_*m*_ represent sensitivities of PCR and multiplex tests, respectively; *T*_*pcr*_ and *T*_*m*_ represent the number of symptomatic PCR SARS-CoV-2 tests on day *t* and the number of symptomatic multiplex PCR tests on day *t*, respectively; and *p(t)* represents the true probability of being infected with virus-2 among tested symptomatic individuals:

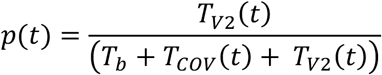

#### S2.4 Sensitivity analyses

##### S2.4.1. Impact of sensitivity of multiplex PCR tests (*s*_*m*_) on the correction of the observed SARS-CoV-2 percent positive

**Figure S1.**
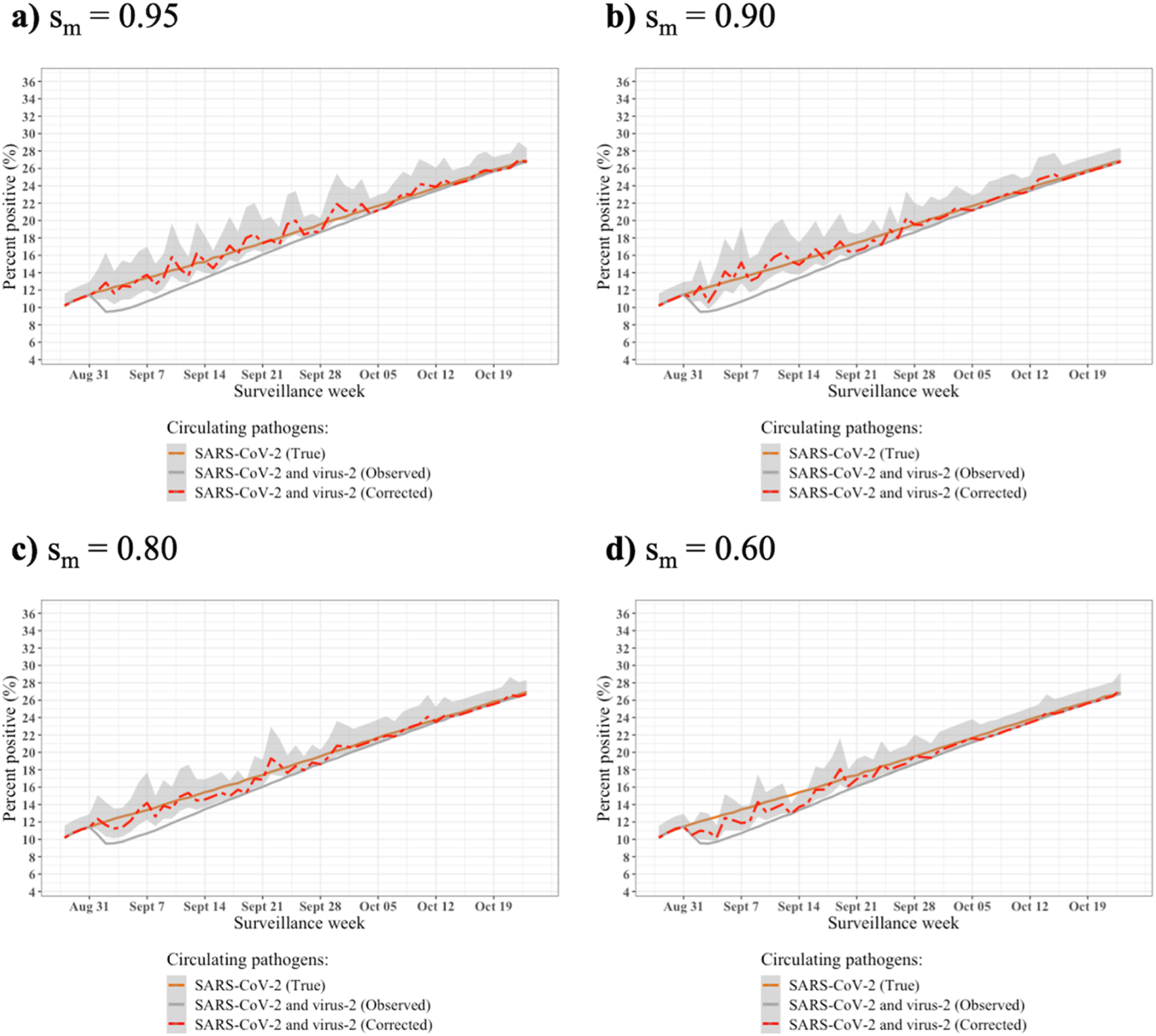
Impact of multiplex PCR test sensitivity (*s*_*m*_) on the correction of the observed SARS-CoV-2 percent positive. Correction of the observed percent positive for SARS-CoV-2 (red dashed line, grey areas represent 95% confidence intervals) depends on the sensitivity of the multiplex PCR assays (s_m_). Figure shows simulations results with a) 0.95, b) 0.90, c) 0.80, and d) 0.60 values of *s*_*m*_. *s*_*m*_ = 0.95 will provide a correction that closely follows real percent positive, while tests with lower *s*_*m*_ provide a correction that is more distant than the real percent positive. For all simulations, we fixed the sensitivity of PCR SARS-CoV-2 test to *s*_*pcr*_ = 0.95 and the proportion of multiplex PCR tests carried out to *m* = 0.001.

##### S2.4.2 Impact of the proportion of multiplex PCR tests used (*m*) among all symptomatic tests on the correction of the observed SARS-CoV-2 percent positive

**Figure S2.**
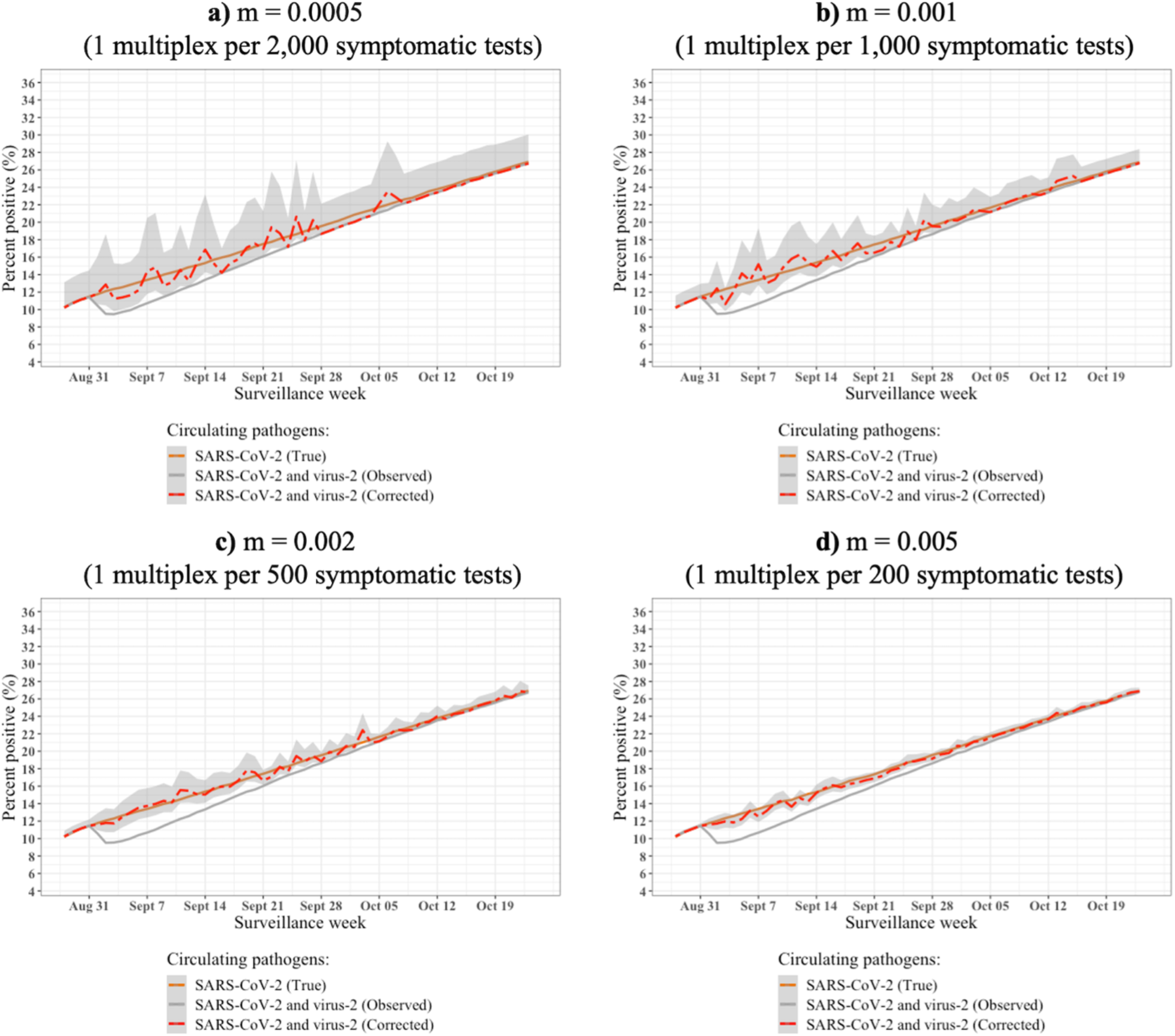
Impact of a range of proportions of multiplex PCR tests used (*m*) among all symptomatic tests on the correction of the observed symptomatic SARS-CoV-2 percent positive with. Quality of the correction of the observed SARS-CoV-2 percent positive (red dashed line, grey areas represent 95% confidence intervals) depends on the number (i.e., proportion) of multiplex PCR tests used (*m*) on symptomatic individuals. Figure shows simulations with a) 0.005, b) 0.001, c) 0.002, and d) 0.005 values of *m*. Greater values of *m* provide better quality of the correction of the observed SARS-CoV-2 percent positive. For all simulations, we fixed the sensitivity of PCR SARS-CoV-2 test to s_pcr_ = 0.95 and the sensitivity of multiplex PCR test to s_m_ = 0.90.

##### S2.4.3 Impact of sensitivity of SARS-CoV-2 PCR tests (*s*_*pcr*_) on the correction of the observed SARS-CoV-2 percent positive

**Figure S3.**
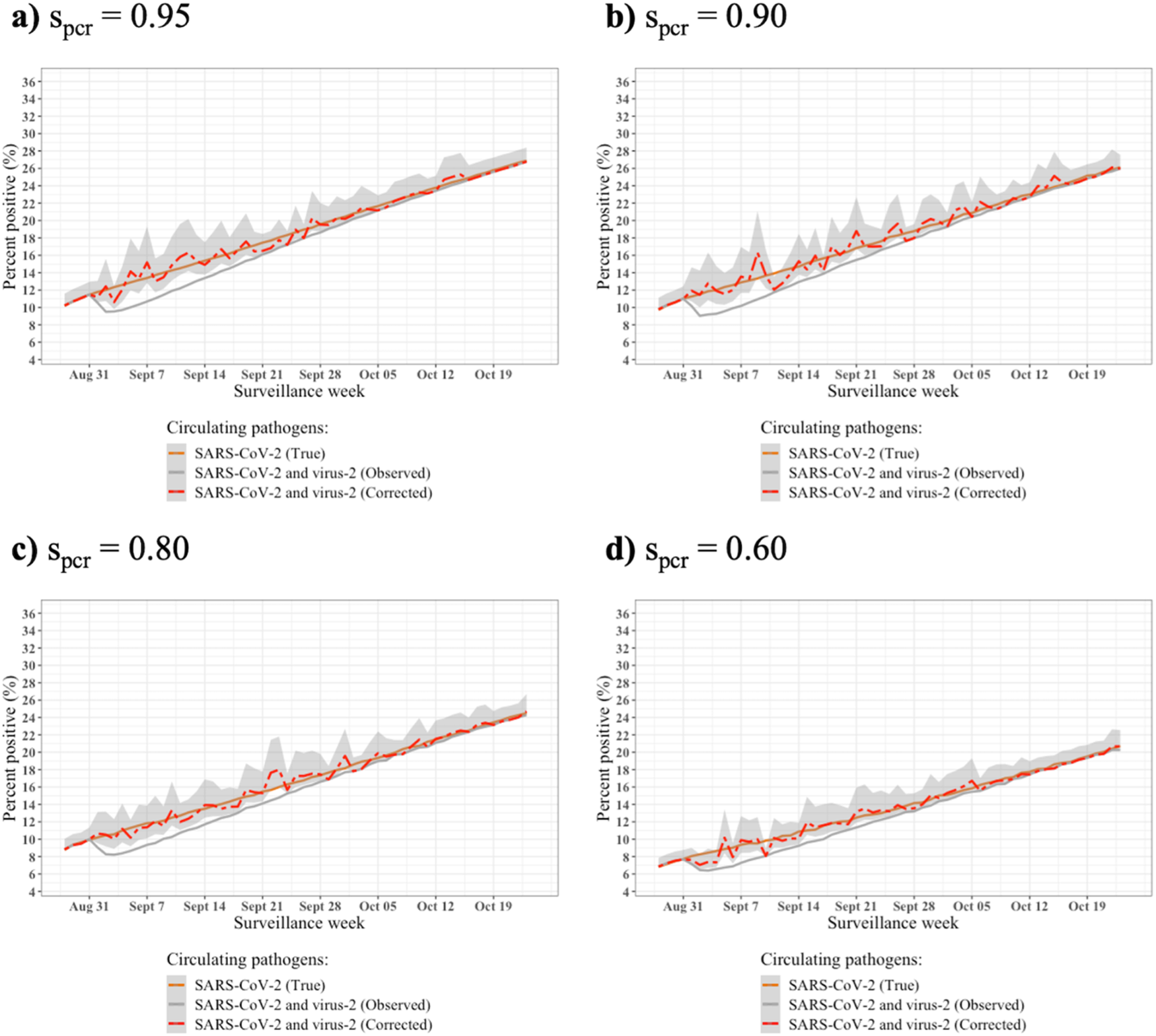
Impact of SARS-CoV-2 PCR test sensitivity (*s*_*pcr*_) on the correction of the observed SARS-CoV-2 percent positive. Correction of the observed percent positive for SARS-CoV-2 (red dashed line, grey areas represent 95% confidence intervals) does not depend on the sensitivity of the SARS-CoV-2 PCR assays (s_pcr_). Figure shows simulations results with a) 0.95, b) 0.90, c) 0.80, and d) 0.60 values of *s*_*pcr*_. Successful detection of positive SARS-CoV-2 cases and overall observed percent positivity depends on *s*_*pcr*_. However, the correction quality is not affected by this parameter. For all simulations, we fixed the sensitivity of multiplex PCR test to *s*_*m*_ = 0.90 and the proportion of multiplex PCR tests carried out to *m* = 0.001.

##### S2.4.4 Impact of the proportion of individuals infected with virus-2 that is symptomatic at the time of testing (*s*_*2*_) on the correction of the observed SARS-CoV-2 percent positive

**Figure S4.**
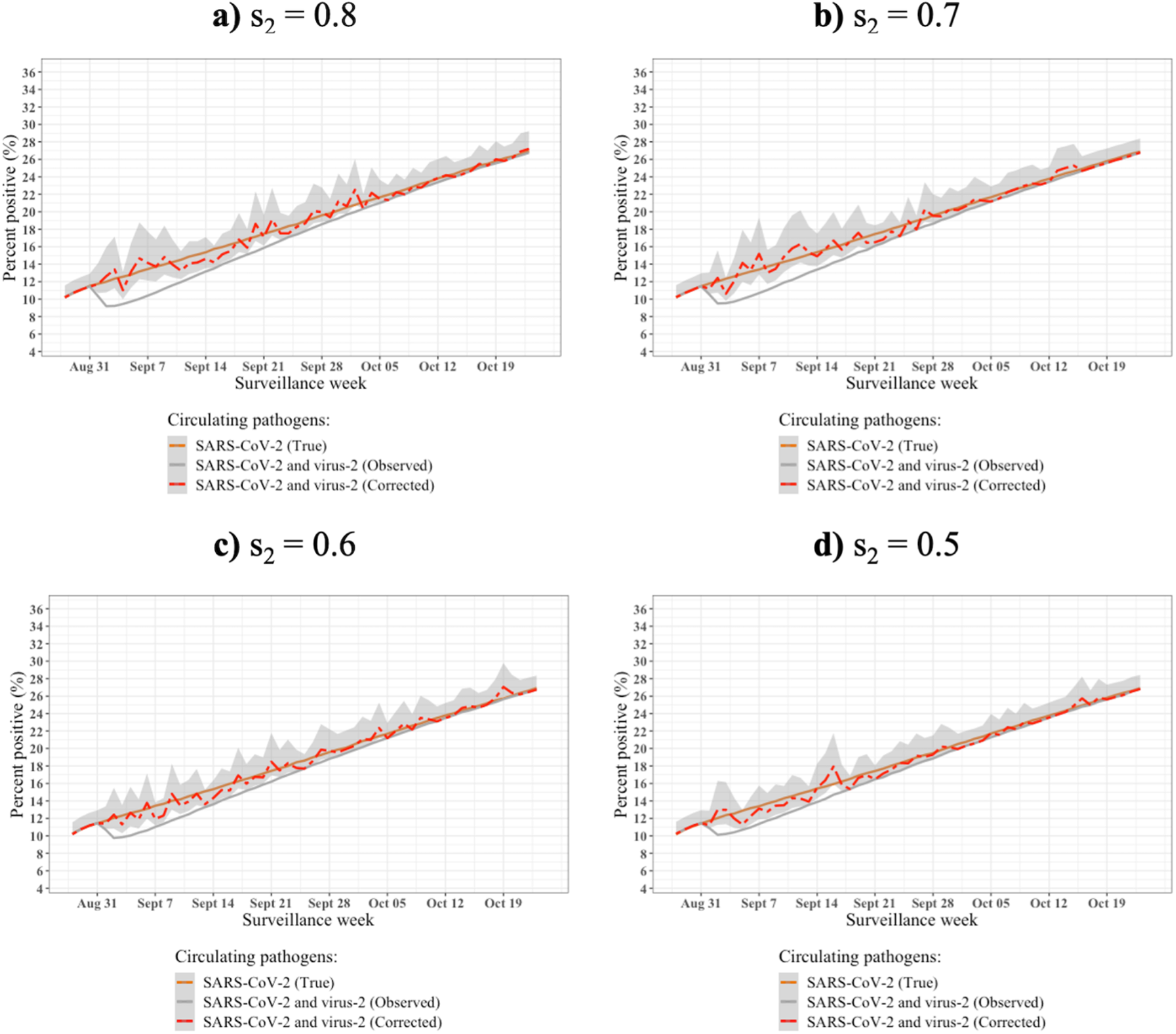
Impact of a range of proportion values for individuals infected with virus-2 that are symptomatic at the time of testing (*s*_*2*_) on the correction of the observed SARS-CoV-2 percent positive. Correction of the observed percent positive for SARS-CoV-2 (red dashed line, grey areas represent 95% confidence intervals) does not depend on the proportion of infected with virus-2 that is symptomatic at the time of testing (*s*_*2*_). Therefore, the correction quality is not affected by this parameter. However, the gap between the true and observed percent positive decreases with the decreasing proportion of infected with virus-2 that is symptomatic due to the fact that less symptomatic individuals infected with virus-2 request SARS-CoV-2 test. Figure shows simulations results with a) 0.8, b) 0.7, c) 0.6, and d) 0.5 values of *s*_*2*_. For all simulations, we fixed the sensitivity of PCR SARS-CoV-2 test to *s*_*pcr*_ = 0.95, sensitivity of multiplex PCR test to *s*_*m*_ = 0.90, and the proportion of multiplex PCR tests carried out to *m* = 0.001.

##### S2.4.5 Impact of virus-2 R_0_ on the correction of the observed SARS-CoV-2 percent positive

**Figure S5.**
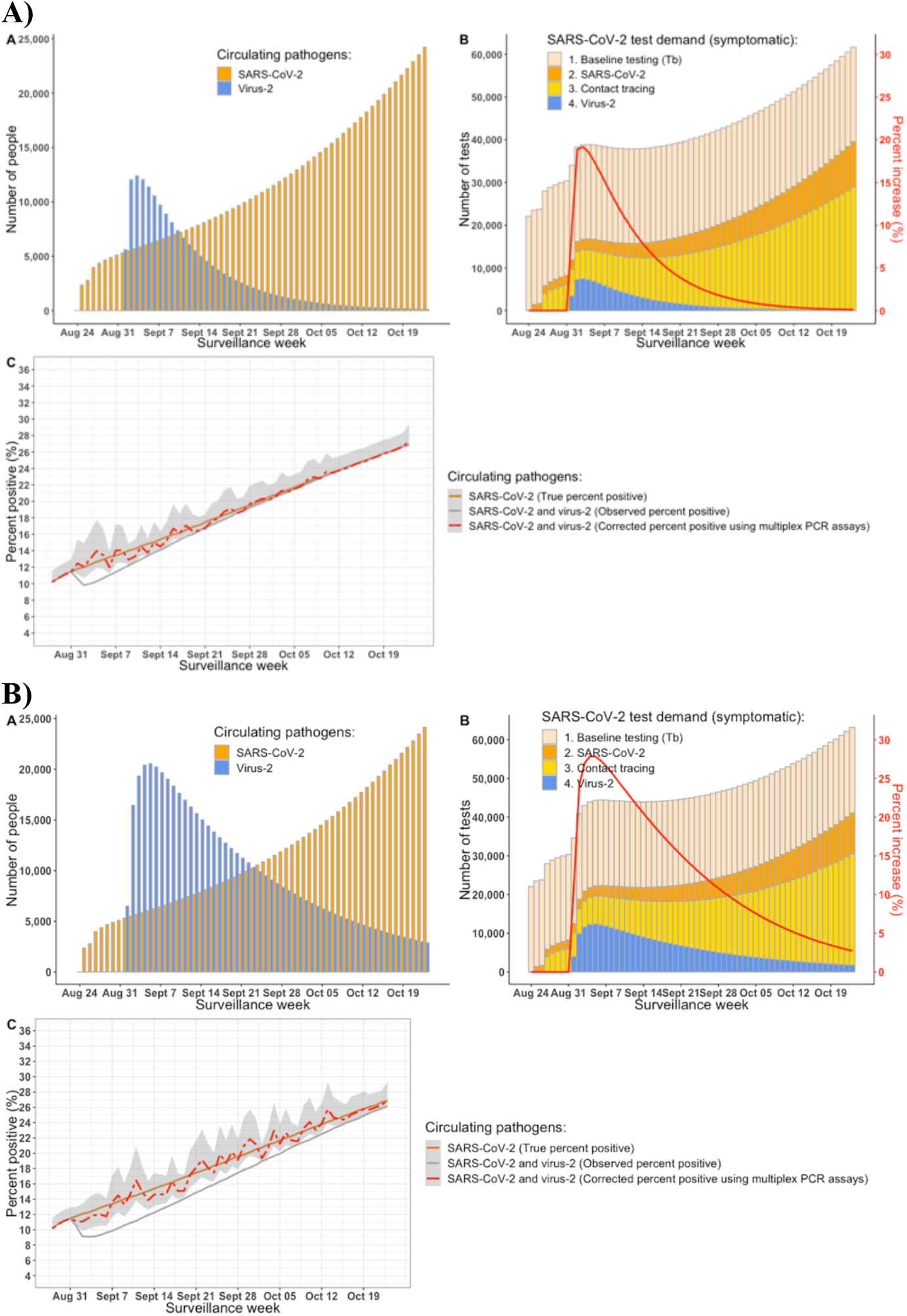
The impact of varying R_0_ of virus-2. Correction of the observed percent positive for SARS-CoV-2 (red dashed line, grey areas represent 95% confidence intervals) does not depend on R_0_ of virus-2 and the correction quality is not affected by this parameter. However, the gap between the true and observed percent positive increases with the increasing value of R_0_ and vice versa. Percent increase in SARS-CoV-2 testing demand (solid red line) is affected in the similar manner. Figure shows simulation results with R_0_ values of **A) 1.4**: The observed SARS-CoV-2 percent positive among symptomatic tests underestimated the true percent positive by up to 2.4% (19% relative decrease). Daily SARS-CoV-2 testing demand increased up to 19.2%; and **B) 2.5**: The observed SARS-CoV-2 percent positive among symptomatic underestimated the true percent positive by up to 3.6% (27.8% relative decrease). Daily SARS-CoV-2 testing demand increased up to 27.9%.

##### S2.4.6 Impact of the initial proportion of the population immune to SARS-CoV-2 on the correction of the observed SARS-CoV-2 percent positive

**Figure S6.**
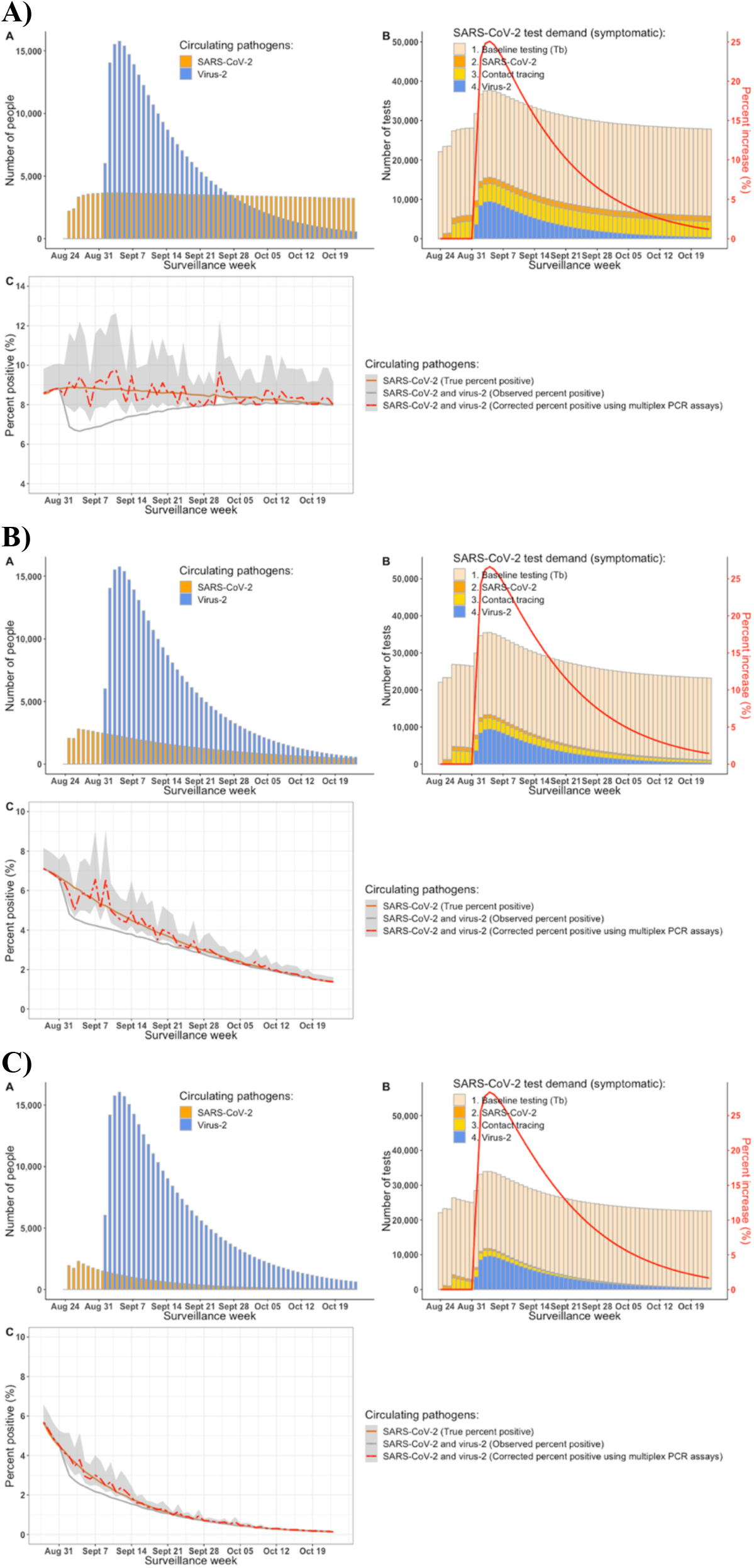
Impact of the initial proportion of the population immune to SARS-CoV-2. Correction of the observed percent positive for SARS-CoV-2 (red dashed line, grey areas represent 95% confidence intervals) does not depend on the initial proportion of the population immune to SARS-CoV-2. The correction quality is not affected by this parameter. However, the gap between the true and observed percent positive decreases with the increasing initial proportion of the population immune to SARS-CoV-2. On the other hand, percent increase in SARS-CoV-2 testing demand (solid red line) experiences slight increase with the increasing initial proportion of the population immune to SARS-CoV-2. Figure shows simulation results with increasing proportion of the population being immune to SARS-CoV-2: **A) 30%** immune to SARS-CoV-2 (*X*_*RS*_ = 0.06; *X*_*SR*_ = 0.46; *X*_*RR*_ = 0.24): The observed SARS-CoV-2 percent positive among symptomatic tests underestimated the true percent positive by up to 2.2% (24.9% relative decrease). Daily SARS-CoV-2 testing demand increased up to 25.1%. **B) 50**% immune to SARS-CoV-2 (*X*_*R*S_ = 0.06; *X*_*SR*_ = 0.26; *X*_*RR*_ = 0.44): The observed SARS-CoV-2 percent positive among symptomatic tests underestimated the true percent positive by up to 1.55% (25.8% relative decrease). Daily SARS-CoV-2 testing demand increased up to 26.6%. **C) 70%** immune to SARS-CoV-2 (*X*_*RS*_ = 0.06; *X*_*SR*_ = 0.06; *X*_*RR*_ = 0.64): The observed SARS-CoV-2 percent positive among symptomatic tests underestimated the true percent positive by up to 0.93% (25.5% relative decrease). Daily SARS-CoV-2 testing demand increased up to 28.4%.

##### S2.4.7 Impact of the initial proportion of the population immune to SARS-CoV-2 and to virus-2 on the correction of the observed SARS-CoV-2 percent positive

**Figure S7.**
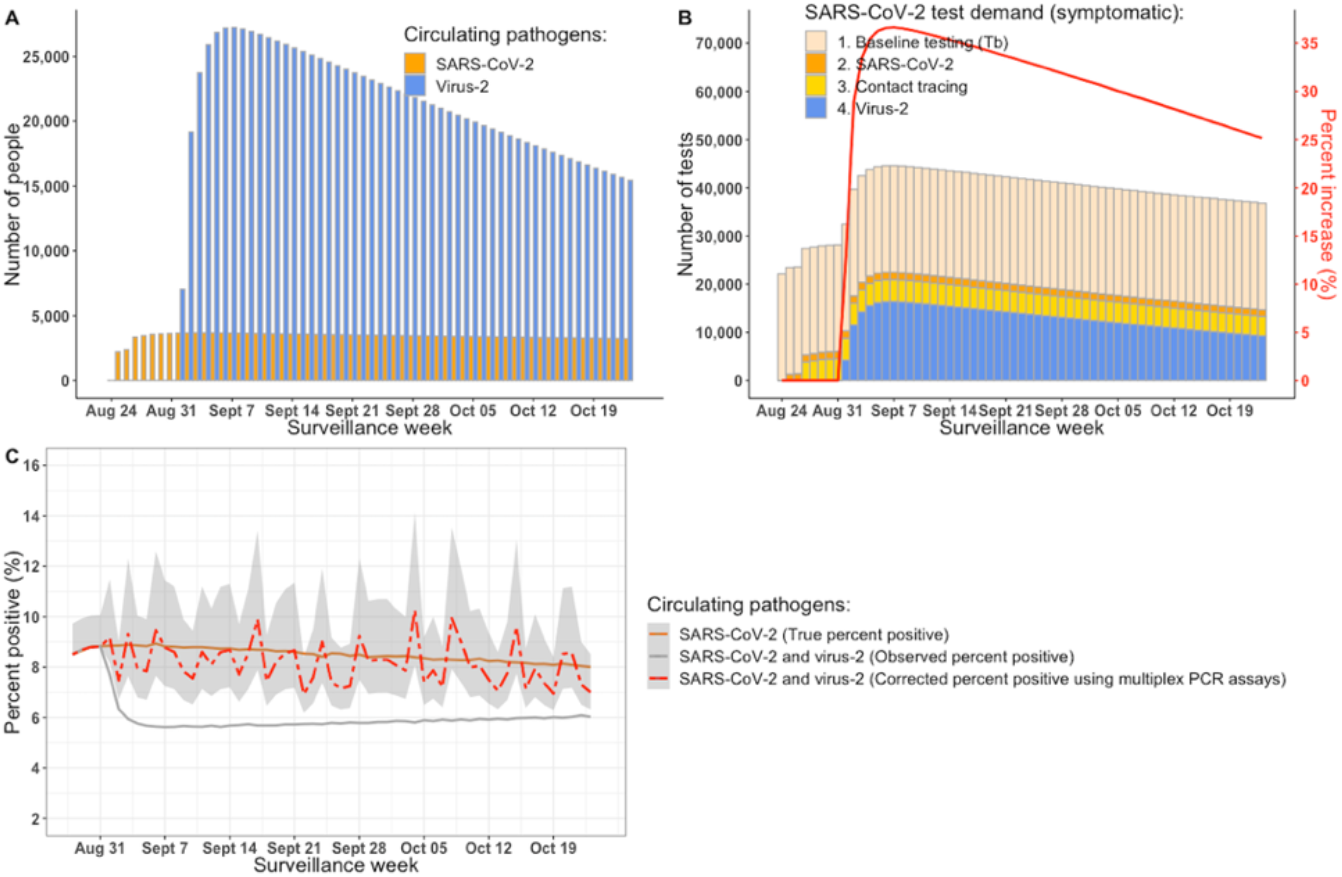
Impact of the initial proportion of the population immune to SARS-CoV-2 and to virus-2. Correction of the observed percent positive for SARS-CoV-2 (red dashed line, grey areas represent 95% confidence intervals) does not depend on the initial proportion of the population immune to SARS-CoV-2. The correction quality is not affected by this parameter. However, both the gap between the true and observed percent positive and percent increase in SARS-CoV-2 testing demand (solid red line). increase with the increasing initial proportion of the population immune to SARS-CoV-2 and decreasing initial proportion of the population immune to virus-2. Figure shows simulation results with **30%** and **50%** of the population being immune to SARS-CoV-2 and virus-2, respectively (*X*_*RS*_ = 0.26; *X*_*SR*_ = 0.46; *X*_*RR*_ = 0.04). The observed SARS-CoV-2 percent positive among symptomatic tests underestimated the true percent positive by up to 3.3% (36.9% relative decrease). Daily SARS-CoV-2 testing demand increased up to 36.6%.

#### S2.5 Rhinovirus circulation in France

**Figure S8.**
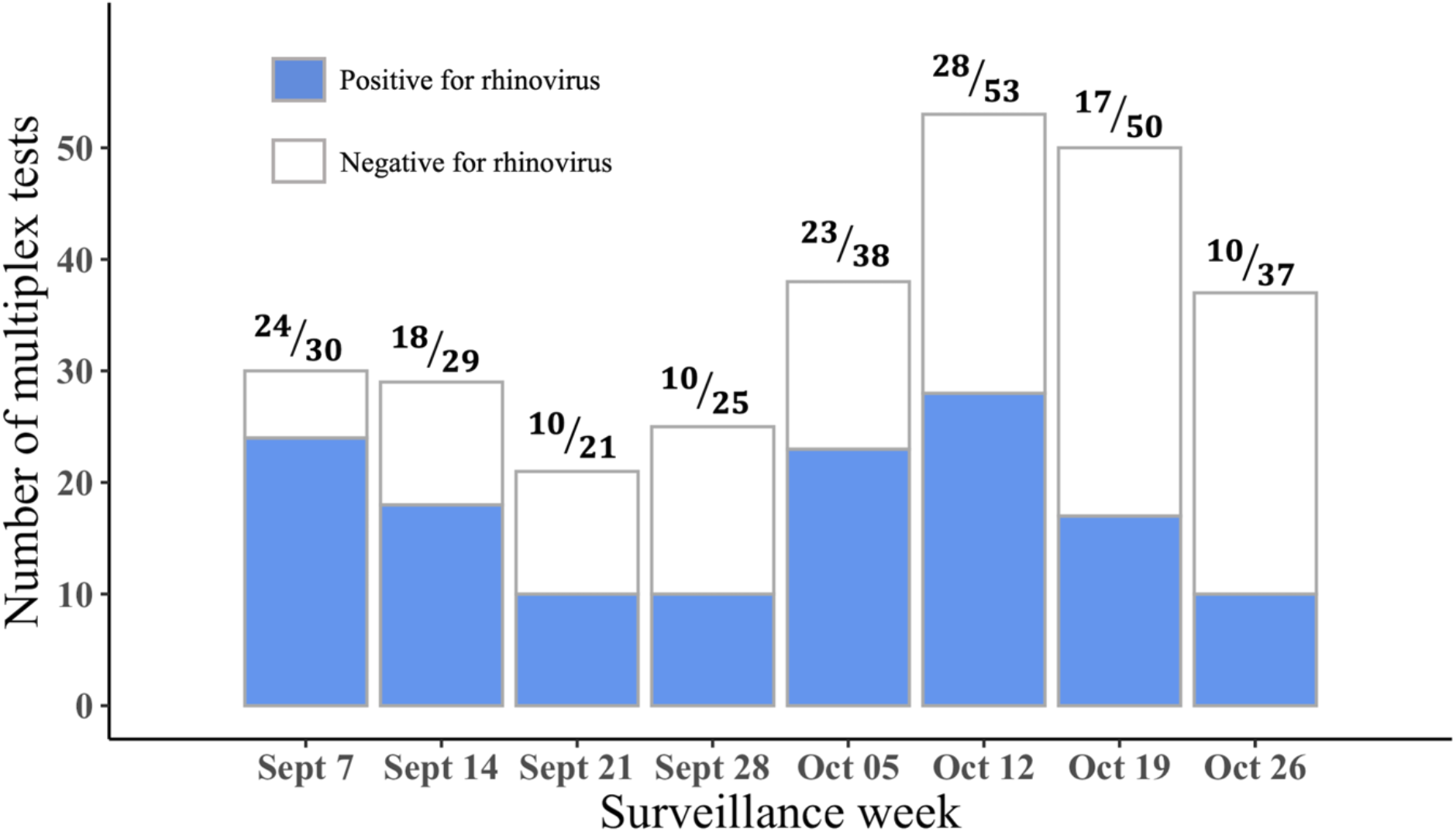
Detection of rhinovirus circulation in France from Sept 7 to Oct 26, 2020. Data are from Ref. [11]. Blue shaded part of the column represents number of multiplex tests positive for rhinovirus (numerator value) out of total number of multiplex tests (denominator value) conducted per week. French Sentinelles physicians (general practitioners and pediatricians) conducted nasopharyngeal swabs among acute respiratory infection (ARI) cases seen in the consultation to test for the various respiratory viruses. This surveillance indicates active circulation of the virus over the study period.

